# Determinants of mortality among COVID-19 patients with diabetes mellitus in Addis Ababa, Ethiopia, 2022: An unmatched case-control study

**DOI:** 10.1101/2022.04.04.22273344

**Authors:** Migbar Sibhat Mekonnen, Melsew Getnet Tsegaw, Wuletaw Chane Zewde, Kassie Gebeyehu Tiruneh, Asaminew Habtamu Sane, Taye Mezgebu Ashine, Hailu Asmare Beyene, Melkie Ambaw Mengistie, Edmialem Getahun Mesfin

## Abstract

**Introduction:** COVID-19 remains one of the leading causes of death seeking global public health attention. Diabetes mellitus is associated with severity and lethal outcomes up to death independent of other comorbidities. Nevertheless, information regarding the determinant factors that contributed to the increased mortality among diabetic COVID-19 patients is limited. Thus, this study aimed at identifying the determinants of mortality among COVID-19 infected diabetic patients.

**Methods:** An unmatched case-control study was conducted on 340 randomly selected patients by reviewing patient records. Data were collected using a structured extraction checklist, entered into Epi data V-4.4.2.2, and analyzed using SPSS V-25. Then, binary logistic regression was used for bivariate and multivariable analysis. Finally, an adjusted odds ratio with 95% CI and a p-value of less than 0.05 was used to determine the strength of association and the presence of a statistical significance consecutively.

**Results:** The study was conducted on 340 COVID-19 patients (114 case and 226 controls). Patient age (AOR=4.90; 95% CI: 2.13, 11.50), severity of COVID-19 disease (AOR=4.95; 95% CI: 2.20, 11.30), obesity (AOR=7.78; 95% CI: 4.05, 14.90), hypertension (AOR=5.01; 95% CI: 2.40, 10.60), anemia at presentation (AOR=2.93; 95% CI: 1.29, 6.65), and AKI after hospital admission (AOR=2.80; 95% CI: 1.39, 5.64) had statistically significant association with increased mortality of diabetic patients with COVID-19 infection. Conversely, presence of RVI co-infection was found to be protective against mortality (AOR=0.35; 95% CI: 0.13, 0.90).

**Conclusion:** Patient age (<65years), COVID-19 disease severity (mild and moderate illness), presence of hypertension, obesity, anemia at admission, and AKI on admission was independently associated with increased mortality of diabetic COVID-19 patients. Contrariwise, the presence of RVI co-infection was found to be protective against patient death. Consequently, COVID-19 patients with diabetes demand untiring efforts, and focused management of the identified factors will substantially worth the survival of diabetic patients infected with COVID-19.

**What is already known on this topic?:** Diabetes mellitus is associated with severity and lethal outcomes up to death independent of other comorbidities. Previous studies indicated that diabetic patients have almost four times increased risk of severe disease and death due to COVID-19 infection. Consequently, with this increased mortality and other public health impacts, numerous reports have been evolved worldwide on the link between COVID-19 and DM, and diabetes management during the COVID-19 pandemic. However, information regarding the determinant factors that lead to the increased mortality among diabetic COVID-19 patients is not well-studied yet.

**What this study adds?:**

- Patient age (<65years), COVID-19 disease severity (mild and moderate illness), presence of hypertension, obesity, anemia at admission, and AKI on hospital admission were independently associated with increased mortality of COVID-19 patients with DM.
- In addition, RVI co-infection was found to be protective against patient death.

## Introduction

The coronavirus disease 2019 (COVID-19) pandemic, which is caused by a highly pathogenic severe acute respiratory syndrome coronavirus 2 (SARS-CoV-2), has resulted in millions of morbidities and mortalities worldwide (1-3). COVID-19 has drawn drastic attention and poses a healthcare threat globally. The devastating effects of the COVID-19 pandemic were highly associated with chronic medical conditions, mainly diabetes mellitus (DM). The estimated global prevalence of diabetes in 2021 was 10.5% (537 million), a 16% (74 million) rise since 2019. The international diabetes federation (IDF) projected that the number of diabetic patients will increase to 11.3% (643 million) by 2030 and 12.2% (783 million) by 2045 (4). The prevalence of diabetes among COVID-19 patients has been estimated up to 31% (5).

Diabetes mellitus (DM) is one of the most frequent comorbidities that occurred in patients with coronavirus disease 2019 (COVID-19) and is associated with severity and lethal outcomes such as intensive care unit (ICU) admission, invasive ventilation, and death (6-8) independent of other comorbidities. Furthermore, the presence of typical complications of diabetes mellitus (cardiovascular disease, heart failure, and chronic kidney disease) increases COVID-19 mortality (6, 9-11). It has been observed that 1.5% of in-hospital deaths related to COVID-19 occurred in patients with type 1 DM and 31.4% in patients with type 2 DM (9).

Deaths in people with diabetes have more than doubled during the COVID-19 epidemic (9). Another study indicates that diabetic patients have almost four times the risk of severe disease and death from COVID-19 (12). The presence of DM is frequently associated with worse outcomes even among patients presented with mild or no COVID-19 symptoms (13).

Consequently, with this increased mortality and other public health impacts, numerous reports have been evolved worldwide on the link between COVID-19 and DM, and diabetes management during the COVID-19 pandemic (14). Predicting worse patient outcomes using poor glycemic control is not an innovation. For instance, fasting blood sugar level was identified as an independent predictor of severity during the 2009 H1N1 influenza pandemic (15). Due to the deregulation of the immune system, diabetic patients are highly vulnerable to infectious diseases. Irrespective of the previous history of DM, hyperglycemia leads to impaired immune function and increased susceptibility to infections, and has been associated with an increased patient death (16, 17).

With the high prevalence of diabetes and its devastating public health consequences, it is demanding to understand the unique aspects of COVID-19 infection in people with diabetes. To control the pandemic, it can be even more significant these days and in the future due to the changes in the lifestyle and healthcare service delivery system. Furthermore, recent studies indicated the need for special considerations in diabetes management among COVID-19 patients (18). Besides, people with diabetes are more concerned about becoming infected with COVID-19 than people without diabetes (19). Thus, this study aimed at identifying the determinants of mortality among COVID-19 infected diabetic patients.

## Methods

An unmatched case-control study was employed by reviewing charts of patients admitted to Addis Ababa COVID-19 care centers from September 2020 to August 2021. Addis Ababa, the capital city of Ethiopia, has three main COVID-19 treatment centers. Millennium COVID-19 care center is the largest and best equipped of all those centers, with a capacity of 1040 patient beds and more than 40 intensive care unit beds. The source population was all COVID-19 patients who died in the center. Patients who died during the specified period were considered the study population. Cases were COVID-19 diagnosed DM patients who died while admitted to the center, whereas controls were COVID-19 patients without a diagnosis of DM and who died in the COVID-19 care center. Medical records without HgA1c and patients with stress hyperglycemia without elevated HgA1c were excluded from the study.

### Sample size and recruitment methods

The sample size was estimated using the double population proportion formula by the statcalc program of Epi info software, considering unmatched case-control study assumptions. The parameters used include 95% confidence level, 80% study power, control to case ratio of two, percentage of controls exposed=60%, and an odds ratio of 2.04 (20) that yields 341. Afterward, 114 cases (diabetic patients) and 228 controls (non-diabetic patients) were incorporated. Cases were selected consecutively, and two respective controls next to the sampled case were included randomly with replacement after confining all death records in their order of medical record number (MRN).

### Data collection tools, procedures, and quality control

Initially, the lists of one-year death records were sorted out. Among those, medical records of diabetic and non-diabetic patients were further confined separately using a computer-generated system and arranged orderly using their medical record number (MRN). Afterward, data collection was conducted using a pretested and structured extraction checklist from medical records of COVID-19 patients who were died in the treatment center. A one-year data (patient records from September 2020 to August 2021) was used to conduct the study. The extraction checklist comprised socio-demographic characteristics, disease severity, comorbidities, baseline vital signs, and laboratory findings, as well as COVID-19 management and complication-related questions. Four data collectors and two supervisors were recruited and trained for half-day about the data collection requirements. The completeness of the collected data was cross-checked daily.

### Statistical analysis and data presentation

After completion of data collection, data were coded and entered into Epi data manager version 4.4.2.2. Then, it was exported to SPSS software version 25 for analysis. Crosstabulation and explorative analysis was done for data cleaning and to determine the data distribution. Categorical data were described using frequency and percent, and mean with standard deviation (SD) or median with interquartile range (IQR) were applied to describe continuous data based on data distribution. Afterward, bivariate and multivariable analyses were run using the binary logistic regression model. After bivariate analysis, a p-value of 0.25 or less was considered to recruit candidate variables for the multivariate model. The model fitness test was checked using Hosmer and Lemeshow test (p=0.302). Finally, an adjusted odds ratio with a 95% confidence interval was used to report the strength of association, and statistical significance was declared at a p-value of less than 0.05.

### Patient and public involvement

It is not applicable for this study since patients were not directly involved. The study was conducted via patient chart review without contacting patients.

### Ethical consideration

Ethical clearance was obtained from Saint Paul’s Millennium Medical College institutional review board (IRB). The IRB waived such that the study can be conducted via patient chart review. Then, the chief executive director and the clinical director were communicated about the purpose of the study, and permission was obtained on behalf of patients to access the data. The study was conducted based on the declaration of Helsinki. Anonymity and confidentiality were attained via coding and aggregate reporting, with no individual patient identifiers collected in the extraction process.

## Results

### Socio-demographic characteristics

This case-control study was conducted on 114 cases (diabetic patients) and 226 controls (non-diabetic patients) that were died due to COVID-19 diseases. About 37.6% of the study subjects were female, of which 27.3% were diabetics. The mean age was 65 years (SD=13.18) among cases and 68.7 years (SD=13.5) among controls ranging from 30 to 94 years. Half of the cases were above the age of 65years, and it was above 62 years among the controls (Table 1).

**Table 1:**
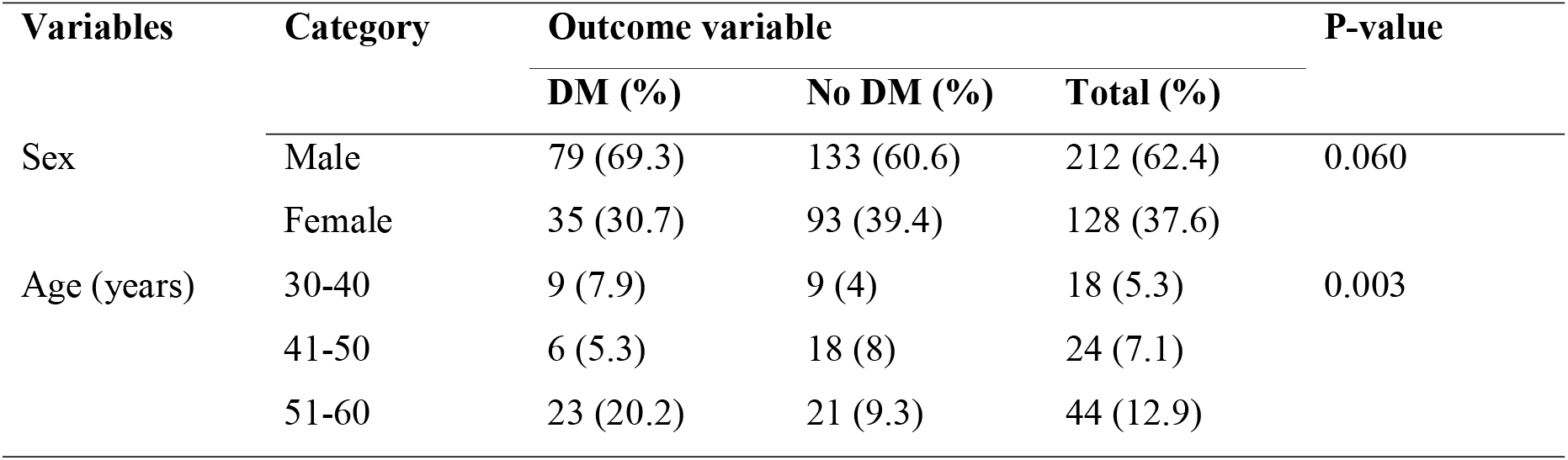
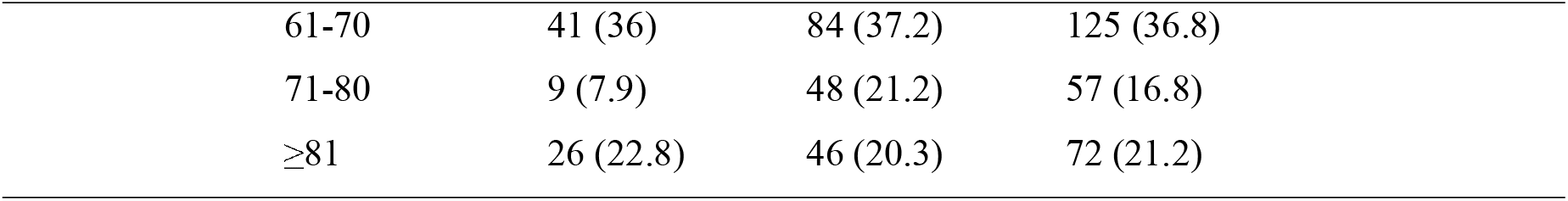
Distribution of socio-demographic characteristics among patients who died due to COVID-19 in Addis Ababa, Ethiopia, 2022 (n=340)

| Variables | Category | Outcome variable |  |  | P-value |
| --- | --- | --- | --- | --- | --- |
|  |  | DM (%) | No DM (%) | Total (%) |  |
| Sex | Male | 79 (69.3) | 133 (60.6) | 212 (62.4) | 0.060 |
|  | Female | 35 (30.7) | 93 (39.4) | 128 (37.6) |  |
| Age (years) | 30-40 | 9 (7.9) | 9 (4) | 18 (5.3) | 0.003 |
|  | 41-50 | 6 (5.3) | 18 (8) | 24 (7.1) |  |
|  | 51-60 | 23 (20.2) | 21 (9.3) | 44 (12.9) |  |

|  |  |  |  |
| --- | --- | --- | --- |
| 61-70 | 41 (36) | 84 (37.2) | 125 (36.8) |
| 71-80 | 9 (7.9) | 48 (21.2) | 57 (16.8) |
| ≥81 | 26 (22.8) | 46 (20.3) | 72 (21.2) |

### COVID-19 severity, comorbidity, and presenting symptoms among patients who died due to COVID-19 patients

The median time from the onset of COVID-19 symptoms to hospital admission was five days (IQR=6) among the cases and seven days (IQR=5) among controls. The study finding also showed that 298 (86.6%) patients were admitted with severe or critical COVID-19 illness, of which 39.3% had diabetes mellitus. The proportion of death among patients admitted with mild and moderate COVID-19 illness was five-fold among the cases (25.4%) compared to non-diabetic patients (5.8%). One-hundred and thirty-seven (40.3%) who died of COVID-19 infection were obese; of those, 58.4% had diabetes mellitus. On the other hand, 82.1% of study participants had comorbid conditions. Hypertension (52.3%), RVI (20.4%), and cardiac illnesses (17.6%) were the commonest comorbidities. Almost two-thirds of the cases (diabetic patients) that died-off COVID-19 diseases were hypertensive, whereas one-third of the controls (non-diabetics) had hypertension. Regarding the presenting COVID-19 symptoms, cough (92.6%) and dyspnea (79.4%) were the frequently reported complaints, followed by easy fatigability (44.4%), myalgia (32.9%), anorexia (16.2%), and chest pain (11.8%) (Table 2).

**Table 2:** COVID-19 severity, comorbidity, and COVID-19 symptoms among patients who died from COVID-19 infection in Addis Ababa, Ethiopia, 2022 (n=340)

| Variable | Category | Outcome variable |  |  | P-value |
| --- | --- | --- | --- | --- | --- |
|  |  | DM (%) | No DM (%) | Total (%) |  |
| COVID-19 Severity | Mild | 5 (4.4) | 1 (0.4) | 6 (1.8) | <0.001 |
|  | Moderate | 24 (21.1)) | 12 (5.3) | 36 (10.6) |  |
|  | Severe/critical | 85 (74.5) | 213 (94.3) | 298 (87.6) |  |
| Obesity | No | 34 (29.8) | 169 (74.8) | 203 (59.7) | <0.001 |
|  | Yes | 80 (70.2) | 57 (25.2) | 137 (40.3) |  |
| Comorbidities | No | 15 (13.2) | 46 (20.4) | 61 (17.9) | 0.103 |
|  | Yes | 99 (86.8) | 180 (79.6) | 279 (82.1) |  |
| HTN | No | 46 (40.4) | 148 (65.5) | 194 (57.1) | <0.001 |
|  | Yes | 68 (59.6) | 78 (34.5) | 146 (42.9) |  |
| Cardiac | No | 92 (80.7) | 199 (88.1) | 291 (85.6) | 0.068 |
|  | Yes | 22 (19.3) | 27 (11.9) | 49 (14.4) |  |
| Stroke | No | 106 (93) | 208 (92) | 314 (92.4) | 0.756 |
|  | Yes | 8 (7) | 18 (8) | 26 (7.6) |  |
| CKD | No | 111 (97.4) | 220 (97.3) | 331 (97.4) | 0.990 |
|  | Yes | 3 (2.6) | 6 (2.7) | 9 (2.6) |  |
| RVI | No | 105 (92.1) | 178 (78.8) | 283 (83.2) | 0.002 |
|  | Yes | 9 (7.9) | 48 (21.2) | 57 (16.8) |  |
| Cancer | No | 111 (97.4) | 224 (99.1) | 335 (98.5) | 0.318 |
|  | Yes | 3 (2.6) | 2 (0.9) | 5 (1.5) |  |
| Presenting symptoms | Anorexia | 3 (2.6) | 52 (23) | 55 (16.2) | <0.001 |
|  | Fatigue | 42 (36.8) | 109 (48.2) | 151 (44.4) | 0.046 |
|  | Headache | 9 (7.9) | 27 (11.9) | 36 (10.6) | 0.252 |
|  | Fever | 8 (7) | 45 (19.9) | 53 (15.6) | 0.002 |
|  | Cough | 108 (94.7) | 207 (91.6) | 315 (92.6) | 0.294 |
|  | Dyspnea | 95 (83.3) | 175 (77.4) | 270 (79.4) | 0.204 |
|  | Sore throat | 3 (2.6) | 18 (8) | 21 (6.2) | 0.054 |
|  | Myalgia | 34 (29.8) | 78 (34.5) | 112 (32.9) | 0.385 |
|  | Chest pain | 16 (14) | 24 (10.6) | 40 (11.8) | 0.356 |

**Key:** DM: diabetes mellitus, COVID-19: coronavirus disease 2019, HTN: hypertension, CKD: chronic kidney disease, RVI: retroviral infection

**Table 3:**
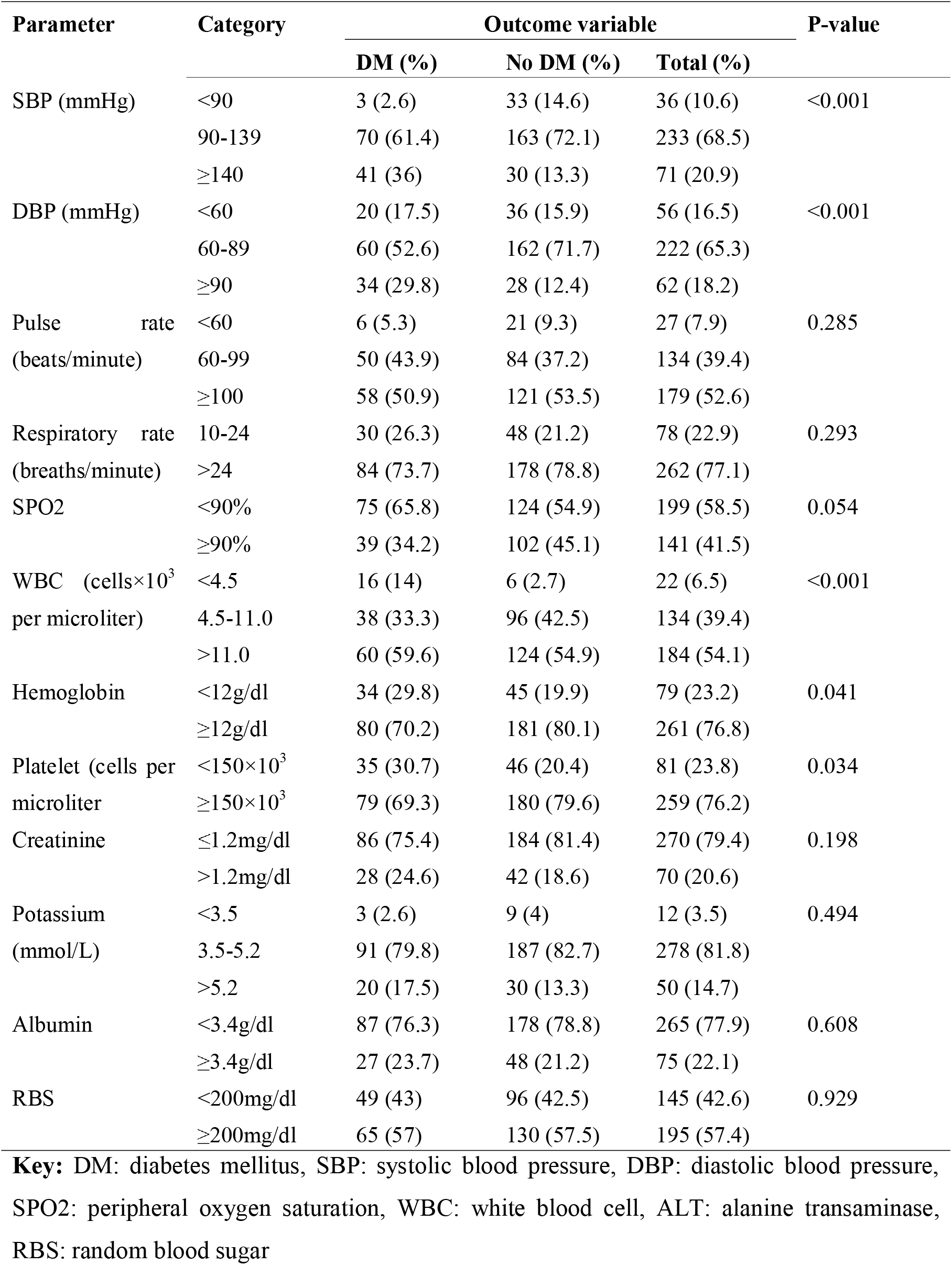
Distribution of baseline vital signs and laboratory investigations among patients who died due to COVID-19 in Addis Ababa, Ethiopia, 2022 (n=340)

| Parameter | Category | Outcome variable |  |  | P-value |
| --- | --- | --- | --- | --- | --- |
|  |  | DM (%) | No DM (%) | Total (%) |  |
| SBP (mmHg) | <90 | 3 (2.6) | 33 (14.6) | 36 (10.6) | <0.001 |
|  | 90-139 | 70 (61.4) | 163 (72.1) | 233 (68.5) |  |
|  | ≥140 | 41 (36) | 30 (13.3) | 71 (20.9) |  |
| DBP (mmHg) | <60 | 20 (17.5) | 36 (15.9) | 56 (16.5) | <0.001 |
|  | 60-89 | 60 (52.6) | 162 (71.7) | 222 (65.3) |  |
|  | ≥90 | 34 (29.8) | 28 (12.4) | 62 (18.2) |  |
| Pulse rate<br>(beats/minute) | <60 | 6 (5.3) | 21 (9.3) | 27 (7.9) | 0.285 |
|  | 60-99 | 50 (43.9) | 84 (37.2) | 134 (39.4) |  |
|  | ≥100 | 58 (50.9) | 121 (53.5) | 179 (52.6) |  |
| Respiratory rate<br>(breaths/minute) | 10-24 | 30 (26.3) | 48 (21.2) | 78 (22.9) | 0.293 |
|  | >24 | 84 (73.7) | 178 (78.8) | 262 (77.1) |  |
| SPO2 | <90% | 75 (65.8) | 124 (54.9) | 199 (58.5) | 0.054 |
|  | ≥90% | 39 (34.2) | 102 (45.1) | 141 (41.5) |  |
| WBC (cells×10<sup>3</sup><br>per microliter) | <4.5 | 16 (14) | 6 (2.7) | 22 (6.5) | <0.001 |
|  | 4.5-11.0 | 38 (33.3) | 96 (42.5) | 134 (39.4) |  |
|  | >11.0 | 60 (59.6) | 124 (54.9) | 184 (54.1) |  |
| Hemoglobin | <12g/dl | 34 (29.8) | 45 (19.9) | 79 (23.2) | 0.041 |
|  | ≥12g/dl | 80 (70.2) | 181 (80.1) | 261 (76.8) |  |
| Platelet (cells per<br>microliter) | <150×10<sup>3</sup> | 35 (30.7) | 46 (20.4) | 81 (23.8) | 0.034 |
|  | ≥150×10<sup>3</sup> | 79 (69.3) | 180 (79.6) | 259 (76.2) |  |
| Creatinine | ≤1.2mg/dl | 86 (75.4) | 184 (81.4) | 270 (79.4) | 0.198 |
|  | >1.2mg/dl | 28 (24.6) | 42 (18.6) | 70 (20.6) |  |
| Potassium<br>(mmol/L) | <3.5 | 3 (2.6) | 9 (4) | 12 (3.5) | 0.494 |
|  | 3.5-5.2 | 91 (79.8) | 187 (82.7) | 278 (81.8) |  |
|  | >5.2 | 20 (17.5) | 30 (13.3) | 50 (14.7) |  |
| Albumin | <3.4g/dl | 87 (76.3) | 178 (78.8) | 265 (77.9) | 0.608 |
|  | ≥3.4g/dl | 27 (23.7) | 48 (21.2) | 75 (22.1) |  |
| RBS | <200mg/dl | 49 (43) | 96 (42.5) | 145 (42.6) | 0.929 |
|  | ≥200mg/dl | 65 (57) | 130 (57.5) | 195 (57.4) |  |

**Key:** DM: diabetes mellitus, SBP: systolic blood pressure, DBP: diastolic blood pressure, SPO2: peripheral oxygen saturation, WBC: white blood cell, ALT: alanine transaminase, RBS: random blood sugar

**Table 4:** Tabulation of management-related variables and COVID-19 complications among patients who died due to COVID-19 in Addis Ababa, Ethiopia, 2022 (n=340)

| Parameter | Category | Outcome variable |  |  | P-value |
| --- | --- | --- | --- | --- | --- |
|  |  | DM (%) | No DM (%) | Total (%) |  |
| Method of ventilation | NP or FM | 10 (8.8) | 5 (2.2) | 15 (4.4) | <0.001 |
|  | NIPPV | 44 (38.6) | 56 (24.8) | 100 (29.4) |  |
|  | IV | 60 (52.6) | 165 (73) | 225 (66.2) |  |
| Diuretics | No | 90 (78.9) | 194 (85.8) | 284 (83.5) | 0.106 |
|  | Yes | 24 (21.1) | 32 (14.2) | 56 (16.5) |  |
| Antihypertensive administered | No | 59 (51.8) | 163 (72.1) | 222 (65.3) | >0.001 |
|  | Yes | 55 (48.2) | 63 (27.9) | 118 (34.7) |  |
| Statins use | No | 94 (82.5) | 191 (84.5) | 285 (83.8) | 0.627 |
|  | Yes | 20 (17.5) | 35 (15.5) | 55 (16.2) |  |
| Antivirals (remdesivir) | No | 101 (88.6) | 209 (92.5) | 310 (91.2) | 0.234 |
|  | Yes | 13 (11.4) | 17 (7.5) | 30 (8.8) |  |
| Prone | No | 37 (32.5) | 48 (21.2) | 85 (25) | 0.024 |
|  | Yes | 77 (67.5) | 178 (78.8) | 255 (75) |  |
| HAI/VAP | No | 37 (32.5) | 61 (27) | 98 (28.8) | 0.294 |
|  | Yes | 77 (67.5) | 165 (73) | 242 (71.2) |  |
| ICU delirium | No | 101 (88.6) | 211 (93.4) | 312 (91.8) | 0.131 |
|  | Yes | 13 (11.4) | 15 (6.6) | 28 (8.2) |  |
| Coagulopathy | No | 90 (78.9) | 175 (77.4) | 265 (77.9) | 0.751 |
|  | Yes | 24 (21.1) | 51 (22.6) | 75 (22.1) |  |
| AKI | No | 58 (50.9) | 138 (61.1) | 196 (57.6) | 0.073 |
|  | Yes | 56 (49.1) | 88 (38.9) | 144 (42.4) |  |
| Shock | No | 51 (44.7) | 99 (43.8) | 150 (44.1) | 0.870 |
|  | Yes | 63 (55.3) | 127 (56.2) | 190 (55.9) |  |
| ALI | No | 81 (71.1) | 165 (73) | 246 (72.4) | 0.703 |
|  | Yes | 33 (28.9) | 61 (27) | 94 (27.6) |  |
| Length of hospital stay (days) | <7 | 33 (29) | 75 (33.2) | 108 (31.8) | 0.696 |
|  | 7-14 | 60 (52.6) | 109 (48.2) | 169 (57.6) |  |
|  | >14 | 21 (18.4) | 42 (18.6) | 63 (18.5) |  |

**Key:** DM: diabetes mellitus, NP: nasal prong, FM: face mask, NIPPV: non-invasive positive pressure ventilation, IV: invasive ventilation, ART: antiretroviral therapy, ARDS: acute respiratory distress syndrome, HAI: hospital-acquired infection, VAP: ventilator-associated pneumonia, ICU: intensive care unit, AKI: acute kidney injury, ALI: acute liver injury

**Table 5:** Binary logistic regression output for the determinant factors of mortality among diabetic COVID-19 patients in Addis Ababa, Ethiopia, 2022 (n=340)

| Covariates | Category | Outcome |  | Crude OR (95% CI) | Adjusted OR (95% CI) | Sig. |
| --- | --- | --- | --- | --- | --- | --- |
|  |  | DM | No DM |  |  |  |
| Gender | Male | 79 | 133 | 1.58 (0.98, 2.55) | 1.34 (0.64, 2.79) | 0.443 |
|  | Female | 35 | 93 | Reference | Reference |  |
| Age in years | <65 | 49 | 72 | 1.61 (1.03, 2.60) | **4.90 (2.13, 11.50)** | **<0.001\*** |
|  | ≥65 | 65 | 154 | Reference | Reference |  |
| COVID-19 severity | Mild or moderate | 30 | 39 | 1.71 (0.99, 2.95) | **4.95 (2.20, 11.30)** | **<0.001\*** |
|  | Severe | 84 | 187 | Reference | Reference |  |
| Obesity | No | 34 | 169 | Reference | Reference |  |
|  | Yes | 80 | 57 | 6.98 (4.20, 11.5) | **7.78 (4.05, 14.90)** | **<0.001\*** |
| HTN | No | 46 | 148 | Reference | Reference |  |
|  | Yes | 68 | 78 | 2.8 (1.80, 4.50) | **5.01 (2.40, 10.60)** | **<0.001\*** |
| Cardiac | No | 92 | 199 | Reference | Reference |  |
| illnesses | Yes | 22 | 27 | 1.76 (0.95, 3.26) | 1.92 (0.77, 4.80) | 0.162 |
| RVI | No | 105 | 178 | Reference | Reference |  |
|  | Yes | 9 | 48 | 0.32 (0.15, 0.67) | **0.35 (0.13, 0.90)** | **0.030\*** |
| Fever | No | 8 | 45 | Reference | Reference |  |
|  | Yes | 106 | 181 | 0.30 (0.14, 0.67) | 0.44 (0.15, 1.27) | 0.128 |
| SPO2 | <90% | 75 | 124 | 1.6 (0.99, 2.50) | 1.14 (0.58, 2.25) | 0.693 |
|  | ≥90% | 39 | 102 | Reference | Reference |  |
| WBC<br>(cells/μL) | ≤11×10<sup>3</sup> | 54 | 102 | Reference | Reference |  |
|  | >11×10<sup>3</sup> | 60 | 124 | 0.91 (0.58, 1.40) | 1.10 (0.58, 2.12) | 0.767 |
| Hemoglobin | <12g/dl | 34 | 45 | 1.71 (1.02,2.9) | **2.93 (1.29, 6.65)** | **0.010\*** |
|  | ≥12g/dl | 80 | 181 | Reference | Reference |  |
| Platelet<br>(cells/μL) | <150×10<sup>3</sup> | 35 | 46 | 1.73 (1.04,2.9) | 0.85 (0.34, 2.02) | 0.710 |
|  | ≥150×10<sup>3</sup> | 79 | 180 | Reference | Reference |  |
| Urea | <60mg/dl | 79 | 184 | Reference | Reference |  |
|  | ≥60mg/dl | 35 | 42 | 1.94 (1.20,3.30) | 1.15 (0.51, 2.50) | 0.774 |
| Proning | No | 37 | 48 | Reference | Reference |  |
|  | Yes | 77 | 178 | 0.56 (0.34, 0.93) | 0.63(0.29, 1.34) | 0.252 |
| Diuretics<br>use | No | 90 | 194 | Reference | Reference |  |
|  | Yes | 24 | 32 | 1.60 (0.90, 2.90) | 2.2(0.96, 4.96) | 0.064 |
| Antiviral<br>use | No | 101 | 209 | Reference | Reference |  |
|  | Yes | 13 | 17 | 1.59 (0.74, 3.39) | 3.5 (0.94, 10.13) | 0.052 |
| Method of<br>ventilation | NP or FM | 10 | 5 | Reference | Reference |  |
|  | NIPPV | 44 | 56 | 0.39 (0.13, 1.23) | 0.69 (0.13, 3.65) | 0.669 |
|  | IV | 60 | 165 | 0.18 (0.06,0.55) | 0.46 (0.10, 2.25) | 0.338 |
| AKI | No | 58 | 138 | Reference | Reference |  |
|  | Yes | 56 | 88 | 1.5 (0.96, 2.39) | **2.80 (1.39, 5.64)** | **0.004\*** |
| ICU<br>delirium | No | 101 | 211 | Reference | Reference |  |
|  | Yes | 13 | 15 | 1.80 (0.83, 3.95) | 2.01 (0.59, 6.88) | 0.266 |

**Note:** \*statistically significant at 95% confidence level
**Key:** DM: diabetes mellitus, HTN: hypertension, RVI: retroviral infection, SPO2: peripheral oxygen saturation, WBC: white blood cell, NP: nasal prong, FM: face mask, NIPPV: non-invasive positive pressure ventilation, IV: invasive ventilation, AKI: acute kidney injury, ICU: intensive care unit

### Baseline vital signs and laboratory investigations

According to the study results, 36% of the cases and 13.3% of the controls had systolic blood pressure above 140mmHg. Additionally, the proportion of tachycardia and tachypnea was comparable among the cases (51% and 73.7%) and the controls (53.5% and 78.7%) respectively. Likewise, 23.2% and 23.8% of the patients were anemic (Hgb<12mg/dl) and thrombocytopenic (Plt<150) in that order. The average WBC count was 11.14 (SD=5.34) among diabetic patients and 13.65 (SD=5.7) among patients without DM. The mean HgA1c among DM patients was 7.1% (SD=1.80), whereas it was 5.34 (SD=0.99) among non-diabetics. Moreover, 57.4% of the patients had random blood sugar (RBS) level of 200mg/dl at baseline, with a comparable proportion among cases and controls. About 17.5% of the cases and 13.3% of the controls had hyperkalemia (K^+^>5.2mmol/L) at admission. At the time of hospital admission, the cases had a serum albumin level of 2.70 (IQR=1.19), and it was 2.19 (IQR=1.14) among controls. Hypoalbuminemia (<3.4g/dl) was recorded among 76.3% of the cases and 78.8% of the controls on admission.

### Description of management-related variables and complications

The median time from hospital admission to death was ten days (IQR=7) among the cases (diabetic patients) and eight days (IQR=8) among controls (patients without DM). The study findings notified that 95.6% of the study participants received either non-invasive or invasive mechanical ventilator support. Two-thirds (66.7%) of the patients who were supported with nasal prong or face mask were diabetics, and only one-third (33.3%) were without diabetes. All patients took antibiotics, thromboprophylaxis, and corticosteroid therapy. Besides, 48.2%, 21.1%, 17.5%, and 11.4% of the cases received antihypertensive medications, diuretics, cholesterol-lowering drugs, and antivirals such as remdesivir respectively. Concerning the COVID-19 related complications, 49.1% of the cases developed AKI, which is true among 38.9% of the controls.

### Determinants of mortality among COVID-19 patients with diabetes mellitus

After data description, inferential analysis was conducted using the binary logistic regression model. Bivariate analysis was done to determine the association of each explanatory variable to the outcome variable. Then, the multi-collinearity chesk was done, and some variables such as antihypertensive use and pulse rate at baseline were excluded since they have a variance inflation factor (VIF) score of greater than ten. Then, variables that fulfilled the test assumptions and had a p-value of less than 0.25 in the bivariate analysis were candidates for the final model. After checking the model fitness test, a multivariate analysis was run by incorporating the candidate variables. Finally, having obesity, hypertension, retroviral infection, anemia at admission, AKI after hospital admission, COVID-19 severity, and patients’ age were found to have a statistically significant association with mortality of diabetic and COVID-19 infected patients.

## Discussion

COVID-19 disease remains one of the leading causes of death seeking public health attention. The mortality rate is even higher among diabetic patients (21). Several studies conducted in the COVID-19 era also reported a strong association between DM and COVID-19 mortality. Nevertheless, information regarding the determinants factors that lead to the increased mortality among diabetic COVID-19 patients is limited. Thus, this study aimed at identifying the determinants of mortality among COVID-19 infected diabetic patients.

The odds of death among diabetic patients under 65 years of age were almost five-fold [(AOR=4.90; 95% CI: 2.13, 11.50), p=<0.001] higher than non-diabetic patients in the same age group did. Previous studies showed the association of COVID-19 patients with advanced age (>65 years). This finding is evidenced in the general population (diabetic and non-diabetic patients) (22, 23). Conversely, the current study reported that patients younger than 65 years are also at a higher risk of death than those without DM. This association might be rationalized by the immune suppression frequently noted in patients with DM than in the general population. This finding implies the need for similar concerns for all diabetic COVID-19 patients irrespective of age strata.

The study finding also revealed that diabetic patients who were admitted with mild and moderate COVID-19 disease had five times [(AOR=4.95; 95% CI: 2.20, 11.30), p=<0.001] increased chance of mortality compared with non-diabetic patients with the same disease severity. Even though not specific to patients with DM, existing evidence documented high mortality among patients who presented with severe COVID-19 illness (23). Equally, the current study notified that having DM would raise the risk of mortality even among those admitted with mild and moderate COVID-19 illness. The scientific reason might be that SARS-COV-2 infection may worsen the disease for people with diabetes by suppressing β-cell function and exacerbating acute metabolic complications (24). The other possible justification could be the weak host immune defense evidenced among diabetic patients that is incapable to halt the viral replication and progress of infection. Consequently, mild and moderate illnesses can be frequently progressed to severe disease among patients with DM than in those who have no DM.

Furthermore, obese patients (BMI≥30) were 7.78 times more likely to die compared to their counterparts [(AOR=7.78; 95% CI: 4.05, 14.90), p<0.001]. Earlier studies conducted on risk factors of mortality among COVID-19 patients supported this finding (7, 9, 25, 26). The pathologic link between mortality of diabetes COVID-19 patients and obesity can be multifactorial. First, obesity can make oxygenation and ventilation more difficult by compromising lung capacity and reserve (27). It is also documented that obesity is associated with impaired immune function (28, 29).

Likewise, the odds of death were five times higher among diabetic patients with hypertension comorbidity than those without hypertension [(AOR= 5.01; 95% CI: 2.40, 10.60), p<0.001]. Former studies conducted in China and Nigeria also showed the association of hypertension with increased mortality of COVID-19 patients (30-33). The exact pathologic link between hypertension and COVID-19 is unclear yet. Nevertheless, the literature hypothesized a higher affinity of SARS-COV-2 to the angiotensin-converting-enzyme receptor-2 (ACE2), which will accelerate viral binding to the epithelial cells of vital organs such as the lung and cardiac (34, 35), resulting in cytokine accumulation in these organs. This increased cytokine concentration (cytokine storm) can lead to acute lung injury, ARDS, and multiple organ failure (36). Besides, cytokine storm is highly linked with the progression of hypertension (37) which could result in severe inflammatory responses and complications.

Diabetic patients who had anemia at presentation (Hgb<12g/dl) were at advanced risk of mortality compared to those with normal hemoglobin levels [(AOR=2.93; 95% CI: 1.29, 6.65), p=0.010). A prospective study conducted on 1257 Iranian COVID-19 patients reported similar findings (38). However, studies conducted in China (39) and Italy (40) did not confine with the current study results. The possible reason might be that patients with decreased hemoglobin levels have reduced oxygen-carrying capacity that can precipitate type-1 respiratory failure. Moreover, anemia is associated with sympathetic nervous system activation. That, in turn, escalates the metabolic demand and pulmonary capillary leakage, ending up with ARDS (41). Anemia may also further suppress the already compromised immune defense of diabetic patients; and thus, lead to an increased chance of mortality among COVID-19 patients.

Moreover, the risk of mortality of diabetic patients who developed AKI was 2.80 times higher than their counterparts [(AOR=2.80; 95% CI: 1.39, 5.64), p=0.004]. This finding is supported by the results of earlier studies conducted in the USA(42), Korea (43), France (44), and a meta-analysis (45). This scientific link may be explained by the pathophysiologic mechanism of COVID-19 disease itself. The SARS-COV-2 may enter the ACE2 receptor (ACE2 is highly prevalent in the kidney) and cause direct injury to the kidney, increased pro-inflammatory viral cytokines, and thrombotic events. On the other hand, AKI is highly prevalent in critical patients with hemodynamic instabilities, severe ARDS (requiring high PEEP levels of ventilation), decreased cardiac output, hypervolemia, sepsis, and those who received nephrotoxic drugs (44). All those conditions are associated with more severe illness and higher mortality. Conversely, poorly controlled diabetes mellitus is also associated with kidney injury (nephropathy), which will worsen patient outcomes.

Unexpectedly, patients who had RVI co-infection were 65% [(AOR=0.35; 95% CI: 0.13, 0.90), p=0.030] less likely to die-off COVID-19 disease compared to those without RVI co-infection. Contradicting results were documented regarding the effect of HIV status and COVID-19 mortality. Some studies showed a statistically significant association between RVI on increased mortality of COVID-19 patients (46). On the contrary, several studies declared that no significant association exists between these variables (47-49). All RVI patients in this cohort were on HAART. Most of the ART regimens were hypothesized to have a therapeutic effect on COVID-19 infection (50), and some (such as remdesivir) were partially recommended as supportive management modalities for patients with COVID-19 disease (51). Even though remdesivir and other ART drugs are no more recommended for COVID-19 patients by WHO, they were still under investigation in clinical trials and in use by some clinical set-ups. Hence, this might be the possible reason for a better outcome. The variation in the effect and COVID-19 outcome among RVI patients could be due to the variation in the management guideline.

This study has certain limitations. First, it would be nice to run a stratified analysis for each type of DM and differentiate among newly versus known diabetic patients, which is not the case in our study. It did not enable us to see the possible variation in the mortality and factors affecting it among the two groups (newly diagnosed and chronic DM). Second, the retrospective nature of the study would delimit the strength of evidence drawn from the study since data were not collected primarily for research purposes.

## Conclusion

This case-control study identified pertinent factors including socio-demographic, disease severity, comorbidities, and complications of COVID-19 that can determine the mortality among COVID-19 infected diabetic patients. Patient age (<65years), COVID-19 disease severity (mild and moderate illness), presence of hypertension, obesity, anemia at admission, and AKI on hospital admission was independently associated with increased mortality of COVID-19 patients with DM. In addition, RVI co-infection was found to be protective against patient death. Consequently, COVID-19 patients with DM require untiring efforts irrespective of age specification and disease severity at admission. Besides, focused management of hypertension and other identified factors will significantly worth the survival of diabetic patients infected with COVID-19.

## Data Availability

The data owner of this study is the millennium COVID-19 care center. Thus, extra data that support the findings of this study are available from the corresponding author upon reasonable request and can be shared upon legal request.

## Abbreviations

AKI: acute kidney injury
ALI: acute liver injury
ALT: alanine transaminase
ARDS: acute respiratory distress syndrome
CKD: chronic kidney disease
DBP: diastolic blood pressure
DM: diabetes mellitus
FM: face mask
HAART: highly active antiretroviral therapy
HAI: hospital-acquired infection
HTN: hypertension
ICU: intensive care unit
IV: invasive ventilation
NIPPV: non-invasive positive pressure ventilation
NP: nasal prong
RBS: random blood sugar
RVI: retroviral infection
SBP: systolic blood pressure
SPO2: peripheral oxygen saturation
VAP: ventilator-associated pneumonia
WBC: white blood cell.

## Acknowledgments

The authors acknowledged the hospital administrators, data collectors, supervisors, and record officers.

## Authors’ contributions

MSM generated the concept and developed the proposal. MGT, MSM, & WCZ designed the study and developed the data extraction tool. KGT, TMA, AHS, MAM, EGM, WCZ & HAB participated in data collection and supervision. MSM performed the statistical analysis and drafted the manuscript. MGT, MSM, AHS & TMA edited and formatted the manuscript for publication. All the authors read, critically revised, and approved the final version of the manuscript. Finally, all the contributors agreed to be equally accountable for every aspect of the work.

## Consent for publication

Not applicable

